# Predicting In-hospital Mortality Using Elixhauser Comorbidity in Patients Underwent Single and Multiple Coronary Artery Bypass Surgery

**DOI:** 10.1101/2023.04.02.23288050

**Authors:** Renxi Li

## Abstract

**Background:** Coronary Artery Bypass Graft (CABG) surgery is a high-risk surgery (mortality rate between 2-4 percent) performed in patients with ischemic heart disease (IHD). Cardiovascular diseases are strongly associated with comorbidities. This study aimed to assess the prediction of in-hospital mortality by comorbidities in patients who underwent single, double, triple, or quadruple and more CABG.

**Methods:** The National Inpatient Sample (NIS) database was used to extract patients who received 1, 2, 3, or 4+ CABG between the last quarter of 2015 and 2020. Best-fit model by logistic regressions was used to predict in-hospital mortality by Elixhauser Comorbidity Index (ECI). Moreover, age was adjusted in ECI prediction.

**Results:** In-hospital mortality was best predicted by ECI in patients who underwent 3 CABG (*c*-statistic = 0.6325, 95% CI = 0.6151-0.6499), followed by 4+ CABG (*c*-statistic = 0.6308, 95% CI = 0.5997-0.6620), 1 CABG (*c*-statistic = 0.6194, 95% CI = 0.6108-0.6281), and 2 CABG (*c*-statistic = 0.6193, 95% CI = 0.6069-0.6317). After adjusting for age, ECI adequately predicted in-hospital mortality in 4+ CABG (*c*-statistic = 0.7196, 95% CI = 0.6916-0.7477) and 3 CABG (*c*-statistic = 0.6937, 95% CI = 0.6774-0.7101). The predictive power for age-adjusted ECI was comparable in 1 CABG (*c*-statistic = 0.6674, 95% CI = 0.6594-0.6755) and 2 CABG (*c*-statistic = 0.6664, 95% CI = 0.6549-0.6780).

**Conclusions:** This study showed that ECI alone was a moderate (*c*-statistic 0.6-0.7) predictor of in-hospital mortality in all CABG. Age-adjusted ECI could effectively predict in-hospital mortality, especially in patients who underwent 3 and 4+ CABG.

## Introduction

Coronary Artery Bypass Graft (CABG) surgery is a frequently performed cardiac surgery procedure to relieve symptoms and reduce mortality risk in patients suffering from ischemic heart disease (IHD) ^1–3^. CABG is a high-risk surgery with a crude in-hospital mortality rate between 2-4 percent compared to 0.7% of all surgeries ^1,4–7^. When there is more than one blockage in the coronary arteries, multiple bypasses are needed. Multiple bypasses have been shown to have better long-term outcomes in low-risk patients while high-risk patients have worse peri-operative outcomes ^8^. However, in-hospital mortality differences in single and multiple bypasses have not been assessed. Age has been well-established as an independent predictor for morality in CABG; increased age is associated with increased risk for complications post-CABG ^9–14^. Thus, age, as a high-risk factor, needs to be accounted for when examining the in-hospital mortality difference in single and multiple CABG.

To adjust for risk when evaluating clinical outcomes, it is often necessary to take into account comorbidities ^15,16^. The Elixhauser Comorbidity measure was created to identify a broad range of comorbidities using ICD codes in large-scale administrative data ^17^. In the International Classification of Diseases, Tenth Revision, Clinical Modification (ICD-10-CM) system, there are 38 comorbidities identified ^18^. Many studies have demonstrated the effectiveness of the Elixhauser Comorbidity measure in predicting the risk of in-hospital mortality ^19–21^.

More recently, the Elixhauser Comorbidity measure has been developed into the Elixhauser Comorbidity Index (ECI), which is a weighted scoring system that predicts in-hospital mortality ^22^. During the validation phase of its development, the Elixhauser Comorbidity Index (ECI) demonstrated strong predictive ability (*c*-statistic = 0.777, 95% CI = 0.776-0.778) for in-hospital mortality across all subjects ^22^. Although the ECI has been validated in several disease categories, including several cardiac diseases with high mortality rates such as acute myocardial infarction and non-hypertensive congestive heart failure, it has not been specifically evaluated for predicting in-hospital mortality in patients who underwent CABG procedure ^22^.

This study aimed to assess the prediction of in-hospital mortality by ECI in patients who underwent single, double, triple, or quadruple and more CABG. This study used the National (Nationwide) Inpatient Sample (NIS) database, which is the largest in-patient database that accounts for 20% of discharges in the United States ^23^. In addition, age was adjusted when examining the ECI prediction of in-hospital mortality in patients who underwent CABG.

## Materials and Methods

The NIS database was used to extract patients who received 1, 2, 3, or 4+ CABG using the International Classification of Diseases, Tenth Revision, Procedure Coding System (ICD-10-PCS) between the last quarter of 2015 and 2020. The ICD-10-PCS codes to extract cases were listed in Supplement Table 1. Patient age, the record of in-hospital mortality, and ICD-10-CM were extracted. Elixhauser Comorbidity Software was used to identify 38 Elixhauser Comorbidity to calculate ECI ^24^. The weights for the Elixhauser Comorbidities in ECI calculation were listed in Table 1.

**Table 1.** Characteristics of patients who underwent 1, 2, 3, or 4+ Coronary Artery Bypass Surgery (CABG) from the last quarter of 2015 to 2020 were identified in the National (Nationwide) Inpatient Sample (NIS) database. ***Abbreviations:*** CABG, Coronary Artery Bypass Surgery; ECI, Elixhauser Comorbidities Index; SD, standard deviation.

|  | **Total Case** | **Age, mean ± SD** | **In-hospital Mortality,**<br>**n (%)** | **ECI, mean ± SD**<br>**(min, max)** |
| --- | --- | --- | --- | --- |
| 1 CABG | 190524 | 66.18 ± 10.08 | 4302 (2.26%) | -2.02 ± 5.34 (-30, 39) |
| 2 CABG | 83725 | 66.47 ± 9.95 | 1970 (2.35%) | -2.02 ± 5.41 (-27, 35) |
| 3 CABG | 48147 | 66.15 ± 9.74 | 1017 (2.11%) | -1.99 ± 5.24 (-28, 32) |
| 4+ CABG | 13540 | 65.63 ± 9.76 | 309 (2.28%) | -2.01 ± 5.18 (-25, 30) |

Best-fit model by logistic regressions was used to predict in-hospital mortality by ECI in patients who underwent 1, 2, 3, and 4+ CABG, respectively. Multivariable logistic regressions adjusting for age were performed to assess in-hospital mortality prediction by ECI in a best-fit prediction model in 1, 2, 3, and 4+ CABG, respectively. The receiver operating characteristic (ROC) for each best-fit model was plotted and the area under the ROC curve (AUC) was estimated including a 95% confidence interval (CI).

Each best-fit model’s ability to discriminate in-hospital mortality was characterized by *c*-statistic. When *c*-statistic = 0.5, the model does not predict better than chance, and when *c*-statistic = 1, the model accurately predicts all occurrences of in-hospital death. A *c*-statistic between 0.6 and 0,7 has moderate predictor power. The threshold for *c*-statistic is 0.7 for adequate predictor power.

All analyses were performed by SAS (version 9.4). The study was retrospective and was based on the open NIS database. Thus, this study was IRB-exempted. The author had full access to all the data in the study and takes responsibility for its integrity and the data analysis.

## Results

There were 190524, 83725, 48147, and 13540 patients who underwent 1, 2, 3, and 4+ CABG, respectively, between the last quarter of 2015 and 2020 from NIS. The characteristics of the patients were summarized in Table 1. Patients who underwent 1, 2, 3, and 4+ CABG had a mean age of 66.18 ± 10.08, 66.47 ± 9.95, 66.15 ± 9.74, and 65.63 ± 9.76, respectively. The in-hospital mortality was highest in 2 CABG (1970/83725, 2.35%), followed by 4+ CABG (309/13540, 2.28%), 1 CABG (4302/190524, 2.26%), and 3 CABG (1017/48147, 2.11%). Patients who underwent 1, 2, 3, and 4+ CABG had an average ECI of -2.02 ± 5.34 (min = -30, max = 39), -2.02 ± 5.41 (min = -27, max = 35), -1.99 ± 5.24 (min = -28, max = 32), and -2.01 ± 5.18 (min = -25, max = 30), respectively.

A summary of the identified Elixhauser Comorbidities in patients who underwent 1, 2, 3, and 4+ CABG was presented in Table 2. The most common Elixhauser Comorbidities were uncomplicated hypertension (1 CABG, 50.68%; 2 CABG, 50.59%; 3 CABG, 51.01%; 4+ CABG, 50.30%), complicated hypertension (1 CABG, 36.67%; 2 CABG, 37.35%; 3 CABG, 36.90%; 4+ CABG, 36.90%), diabetes with chronic complications (1 CABG, 31.42%; 2 CABG, 32.64%; 3 CABG, 32.65%; 4+ CABG, 32.65%), obesity (1 CABG, 28.55%; 2 CABG, 28.82%; 3 CABG, 27.94%; 4+ CABG, 27.94%), chronic pulmonary disease (1 CABG, 21.75%; 2 CABG, 21.98%; 3 CABG, 20.89%; 4+ CABG, 20.89%), diabetes without chronic complications (1 CABG, 16.49%; 2 CABG, 16.37%; 3 CABG, 17.69%; 4+ CABG, 17.69%), peripheral vascular disease (1 CABG, 15.06%; 2 CABG, 15.21%; 3 CABG, 14.27%; 4+ CABG, 14.27%), hypothyroidism (1 CABG, 11.16%; 2 CABG, 11.34%; 3 CABG, 10.55%; 4+ CABG, 10.55%), depression (1 CABG, 8.73%; 2 CABG, 8.83%; 3 CABG, 8.19%; 4+ CABG, 8.19%), alcohol abuse (1 CABG, 3.36%; 2 CABG, 3.46%; 3 CABG, 3.43%; 4+ CABG, 3.43%), autoimmune conditions (1 CABG, 2.58%; 2 CABG, 2.56%; 3 CABG, 2.33%; 4+ CABG, 2.33%), severe renal failure (1 CABG, 2.37%; 2 CABG, 2.45%; 3 CABG, 2.28%; 4+ CABG, 2.28%), cerebrovascular disease (1 CABG, 1.97%; 2 CABG, 2.07%; 3 CABG, 2.02%; 4+ CABG, 2.02%), drug abuse (1 CABG, 1.70%; 2 CABG, 1.62%; 3 CABG, 1.67%; 4+ CABG, 1.67%), paralysis (1 CABG, 1.41%; 2 CABG, 1.50%; 3 CABG, 1.45%; 4+ CABG, 1.45%), malignant solid tumor without metastasis(1 CABG, 1.32%; 2 CABG, 1.37%; 3 CABG, 1.22%; 4+ CABG, 1.22%), valvular disease (1 CABG, 1.20%; 2 CABG, 0.86%; 3 CABG, 0.56%; 4+ CABG, 0.56%), dementia (1 CABG, 1.10%; 2 CABG, 1.17%; 3 CABG, 1.13%; 4+ CABG, 1.13%), other thyroid disorders (1 CABG, 1.05%; 2 CABG, 1.05%; 3 CABG, 0.96%; 4+ CABG, 0.96%), leukemia (1 CABG, 0.42%; 2 CABG, 0.41%; 3 CABG, 0.48%; 4+ CABG, 0.48%), lymphoma (1 CABG, 0.37%; 2 CABG, 0.37%; 3 CABG, 0.34%; 4+ CABG, 0.34%), acquired immune deficiency syndrome (1 CABG, 0.36%; 2 CABG, 0.36%; 3 CABG, 0.34%; 4+ CABG, 0.34%), metastatic cancer (1 CABG, 0.23%; 2 CABG, 0.26%; 3 CABG, 0.20%; 4+ CABG, 0.20%), moderate to severe liver disease (1 CABG, 0.09%; 2 CABG, 0.09%; 3 CABG, 0.06%; 4+ CABG, 0.06%), heart failure (1 CABG, 0.08%; 2 CABG, 0.09%; 3 CABG, 0.08%; 4+ CABG, 0.08%), in situ solid tumor without metastasis (1 CABG, 0.02%; 2 CABG, 0.02%; 3 CABG, 0.02%; 4+ CABG, 0.02%). In neither CABG group, there were no cases of anemias due to other nutritional deficiencies, chronic blood loss anemia (iron deficiency), coagulopathy, mild liver disease, neurological disorders affecting movement, other neurological disorders, seizures and epilepsy, psychoses, pulmonary circulation disease, moderate renal failure, peptic ulcer disease x bleeding, or weight loss.

**Table 2.** Elixhauser Comorbidities identified by ICD-10 Clinical Modification (ICD-10-CM) to derive the Elixhauser Comorbidities Index (ECI) in patients who underwent 1, 2, 3, or 4+ Coronary Artery Bypass Surgery (CABG) from the last quarter of 2015 to 2020 in National (Nationwide) Inpatient Sample (NIS) database.

| **Elixhauser Comorbidities** | **1 CABG, n (%)** | **2 CABG, n (%)** | **3 CABG, n (%)** | **4+ CABG, n (%)** | **Weight** |
| --- | --- | --- | --- | --- | --- |
| Acquired immune deficiency syndrome | 690 (0.36%) | 304 (0.36%) | 165 (0.34%) | 40 (0.30%) | -4 |
| Alcohol abuse | 6398 (3.36%) | 2901 (3.46%) | 1650 (3.43%) | 423 (3.12%) | -1 |
| Anemias due to other nutritional deficiencies | 0 (0.00%) | 0 (0.00%) | 0 (0.00%) | 0 (0.00%) | -3 |
| Autoimmune conditions | 4917 (2.58%) | 2142 (2.56%) | 1120 (2.33%) | 295 (2.18%) | 0 |
| Chronic blood loss anemia (iron deficiency) | 0 (0.00%) | 0 (0.00%) | 0 (0.00%) | 0 (0.00%) | -4 |
| Lymphoma | 704 (0.37%) | 307 (0.37%) | 162 (0.34%) | 54 (0.40%) | 9 |
| Leukemia | 809 (0.42%) | 347 (0.41%) | 232 (0.48%) | 43 (0.32%) | 5 |
| Metastatic cancer | 429 (0.23%) | 220 (0.26%) | 95 (0.20%) | 26 (0.19%) | 22 |
| Solid tumor without metastasis, in situ | 41 (0.02%) | 15 (0.02%) | 12 (0.02%) | 5 (0.04%) | 0 |
| Solid tumor without metastasis, malignant | 2511 (1.32%) | 1147 (1.37%) | 585 (1.22%) | 167 (1.23%) | 10 |
| Cerebrovascular disease | 3760 (1.97%) | 1732 (2.07%) | 974 (2.02%) | 271 (2.00%) | 5 |
| Heart failure | 150 (0.08%) | 75 (0.09%) | 39 (0.08%) | 16 (0.12%) | 14 |
| Coagulopathy | 0 (0.00%) | 0 (0.00%) | 0 (0.00%) | 0 (0.00%) | 14 |
| Dementia | 2089 (1.10%) | 982 (1.17%) | 542 (1.13%) | 125 (0.92%) | 5 |
| Depression | 16628 (8.73%) | 7389 (8.83%) | 3943 (8.19%) | 989 (7.30%) | -8 |
| Diabetes without chronic complications | 31412 (16.49%) | 13704 (16.37%) | 8519 (17.69%) | 2514 (18.57%) | -2 |
| Diabetes with chronic complications | 59869 (31.42%) | 27330 (32.64%) | 15719 (32.65%) | 4439 (32.78%) | 0 |
| Drug abuse | 3238 (1.70%) | 1360 (1.62%) | 802 (1.67%) | 265 (1.96%) | -7 |
| Hypertension, complicated | 69861 (36.67%) | 31269 (37.35%) | 17765 (36.90%) | 4980 (36.78%) | 1 |
| Hypertension, uncomplicated | 96561 (50.68%) | 42360 (50.59%) | 24562 (51.01%) | 6811 (50.30%) | 0 |
| Liver disease, mild | 0 (0.00%) | 0 (0.00%) | 0 (0.00%) | 0 (0.00%) | 2 |
| Liver disease, moderate to severe | 165 (0.09%) | 74 (0.09%) | 27 (0.06%) | 14 (0.10%) | 16 |
| Chronic pulmonary disease | 41430 (21.75%) | 18403 (21.98%) | 10057 (20.89%) | 2555 (18.87%) | 2 |
| Neurological disorders affecting movement | 0 (0.00%) | 0 (0.00%) | 0 (0.00%) | 0 (0.00%) | -1 |
| Other neurological disorders | 0 (0.00%) | 0 (0.00%) | 0 (0.00%) | 0 (0.00%) | 22 |
| Seizures and epilepsy | 0 (0.00%) | 0 (0.00%) | 0 (0.00%) | 0 (0.00%) | 2 |
| Obesity | 54403 (28.55%) | 24126 (28.82%) | 13452 (27.94%) | 3762 (27.78%) | -7 |
| Paralysis | 2681 (1.41%) | 1258 (1.50%) | 696 (1.45%) | 191 (1.41%) | 4 |
| Peripheral vascular disease | 28694 (15.06%) | 12731 (15.21%) | 6872 (14.27%) | 1710 (12.63%) | 3 |
| Psychoses | 0 (0.00%) | 0 (0.00%) | 0 (0.00%) | 0 (0.00%) | -9 |
| Pulmonary circulation disease | 0 (0.00%) | 0 (0.00%) | 0 (0.00%) | 0 (0.00%) | 4 |
| Renal failure, moderate | 0 (0.00%) | 0 (0.00%) | 0 (0.00%) | 0 (0.00%) | 3 |
| Renal failure, severe | 4511 (2.37%) | 2052 (2.45%) | 1096 (2.28%) | 294 (2.17%) | 7 |
| Hypothyroidism | 21266 (11.16%) | 9492 (11.34%) | 5081 (10.55%) | 1350 (9.97%) | -3 |
| Other thyroid disorders | 2000 (1.05%) | 882 (1.05%) | 460 (0.96%) | 118 (0.87%) | -8 |
| Peptic ulcer disease x bleeding | 0 (0.00%) | 0 (0.00%) | 0 (0.00%) | 0 (0.00%) | 0 |
| Valvular disease | 2289 (1.20%) | 721 (0.86%) | 271 (0.56%) | 57 (0.42%) | 0 |
| Weight loss | 0 (0.00%) | 0 (0.00%) | 0 (0.00%) | 0 (0.00%) | 13 |

A summary of ECI prediction of in-hospital mortality was shown in Figure 1 and summarized in Table 3. While all of the *c*-statistics did not reach adequacy (*c*-statistic > 0.7), in-hospital mortality was best predicted by ECI in patients who underwent 3 CABG (*c*-statistic = 0.6325, 95% CI = 0.6151-0.6499), followed by 4+ CABG (*c*-statistic = 0.6308, 95% CI = 0.5997-0.6620). The predictive power of ECI in patients who underwent 1 CABG (*c*-statistic = 0.6194, 95% CI = 0.6108-0.6281) and 2 CABG (*c*-statistic = 0.6193, 95% CI = 0.6069-0.6317) were comparable.

**Figure 1.**
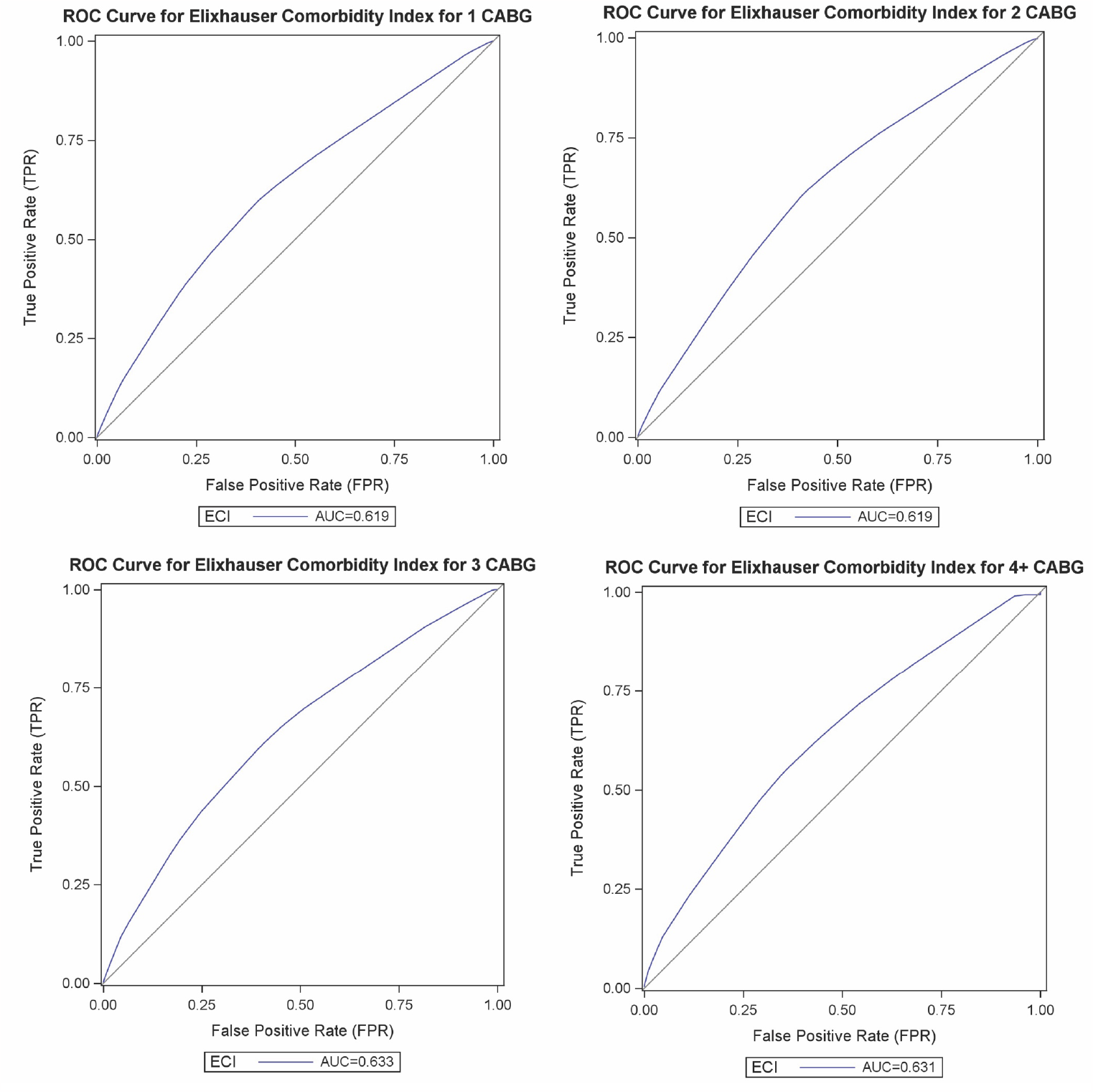
Receiver operating characteristic (ROC) curves for logistic regression models predicting in-hospital mortality by Elixhauser Comorbidities Index (ECI) in patients who underwent 1, 2, 3, or 4+ Coronary Artery Bypass Surgery (CABG) from the last quarter of 2015 to 2020 identified in National (Nationwide) Inpatient Sample (NIS) database. A straight diagonal line shows the null model (AUC 0.500).

**Table 3.** In-hospital mortality discrimination by Elixhauser Comorbidities Index (ECI), as well as ECI, adjusted for age in patients who underwent 1, 2, 3, or 4+ Coronary Artery Bypass Surgery (CABG) from the last quarter of 2015 to 2020 identified in National (Nationwide) Inpatient Sample (NIS) database. ***Abbreviations:*** CABG, Coronary Artery Bypass Surgery; ECI, Elixhauser Comorbidities Index.

|  | **C-Statistic** | **Lower 95% CI** | **Upper 95% CI** |
| --- | --- | --- | --- |
| **ECI** |  |  |  |
| 1 CABG | 0.6194 | 0.6108 | 0.6281 |
| 2 CABG | 0.6193 | 0.6069 | 0.6317 |
| 3 CABG | 0.6325 | 0.6151 | 0.6499 |
| 4+ CABG | 0.6308 | 0.5997 | 0.6620 |
| **ECI adjusted for age** |  |  |  |
| 1 CABG | 0.6674 | 0.6594 | 0.6755 |
| 2 CABG | 0.6664 | 0.6549 | 0.6780 |
| 3 CABG | 0.6937 | 0.6774 | 0.7101 |
| 4+ CABG | 0.7196 | 0.6916 | 0.7477 |

The age-adjusted ECI prediction of in-hospital mortality was also shown in Figure 2 and summarized in Table 3. The predictive power of ECI in all groups of patients was improved by the adjustment of age. The prediction of in-hospital mortality in 4+ CABG patients was adequate (*c*-statistic = 0.7196, 95% CI = 0.6916-0.7477), and that in 3 CABG patients was approaching adequacy (*c*-statistic = 0.6937, 95% CI = 0.6774-0.7101). The predictive power of ECI in patients who underwent 1 CABG (*c*-statistic = 0.6674, 95% CI = 0.6594-0.6755) and 2 CABG (*c*-statistic = 0.6664, 95% CI = 0.6549-0.6780) were still comparable after adjusting for age.

**Figure 2.**
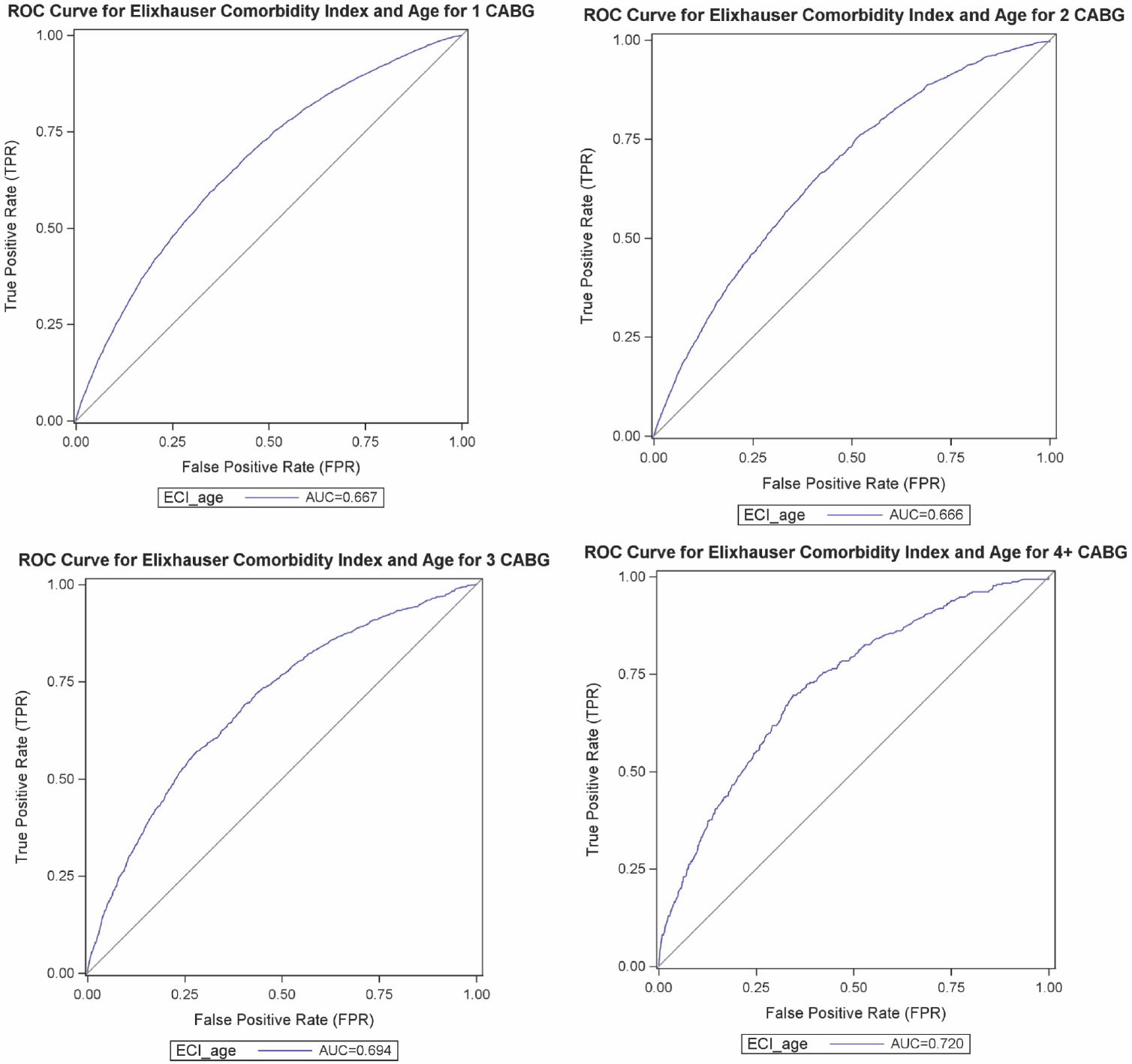
Receiver operating characteristic (ROC) curves for multivariable logistic regression models predicting in-hospital mortality by Elixhauser Comorbidities Index (ECI) after adjusting for age in patients who underwent 1, 2, 3, or 4+ Coronary Artery Bypass Surgery (CABG) from the last quarter of 2015 to 2020 identified in National (Nationwide) Inpatient Sample (NIS) database. A straight diagonal line shows the null model (AUC 0.500).

## Discussion

This study examined the prediction of in-hospital mortality by ECI in patients who underwent 1, 2, 3, and 4+ CABG in NIS data from the last quarter of 2015 to 2020. In addition, age, as a strong risk factor for in-hospital mortality in CABG, was adjusted in the predictive model by ECI.

ECI was found to be a moderate (*c*-statistic > 0.6) predictor for in-hospital mortality in all groups of CABG patients. ECI slightly better discriminated in-hospital mortality in 3 and 4+ (*c*-statistic = 0.6325 and 0.6308, respectively) than 1 and 2 (*c*-statistic = 0.6194 and 0.6193, respectively) CABG patients. In other words, in patients who underwent a high number (>2) of coronary artery bypasses, sicker patients (with more comorbidities) were more associated with mortality. This is clinically justifiable since cardiovascular disease is strongly associated with comorbidities, such as diabetes, hypertension, and dyslipidemia ^25^. Patients with worse overall health conditions are more likely to develop more advanced cardiovascular diseases such as multiple coronary artery blockages that have worse prognoses after the surgery. As a result, comorbidities and mortality are more likely to have an association in patients who undergo multiple coronary artery bypasses.

Although the predictive power of ECI was improved after adjusting for age, the extent of improvement was different depending on the number of coronary bypasses. While the *c*-statistic increased by 0.5-0.6 in patients who underwent 1, 2, and 3 CABG, the *c*-statistic increased by about 0.9 in 4+ CABG patients. The age-adjusted ECI was found to adequately predict (*c*-statistic = 0.7196) in-hospital mortality in 4+ CABG patients, meaning age is a greater risk factor for mortality in these patients. In 3 CABG patients, age-adjusted ECI could also adequately discriminate (*c*-statistic = 0.6937) in-hospital mortality. Thus, age-adjusted ECI is especially effective to predict in-hospital mortality in patients underwent a high number (>2) of coronary artery bypasses.

There were several limitations of this study. First, all in-hospital mortality examined in this study did not have any follow-up in NIS. It was also argued that the 30-day post-surgery period should be monitored for early perioperative outcomes ^26^. Secondly, The Elixhauser Comorbidities were documented in a binary format, indicating whether they were present or absent. This allowed for greater diversity among patients with the same comorbidity, as they may have varying degrees or stages of the disease. In addition, many risk factors in CABG, such as ejection fraction, previous open heart operations, and serum creatine, that can adjust ECI prediction are not recorded in NIS ^27,28^. ECI should be used together with other relevant clinical information, such as specific diagnosis, past medical history, and lab values to accurately assess mortality risk.

Nevertheless, this study showed that age-adjusted ECI can effectively predict in-hospital mortality, especially in patients who underwent 3 and 4+ CABG. Future studies can examine the effect of graft properties, such as venous, arterial, or hybrid grafting, and bypass procedures, such as off-pump versus minimally invasive, on adjusting the prediction of in-hospital mortality by ECI in CABG patients. Furthermore, the weight of the comorbidities in ECI as shown in Table 2 was derived in the larger population. For example, valvular disease was given a weight of 0 in ECI, but the presence of valvular disease might be a predisposition factor for mortality in CABG and thus should be given a positive weight. Future studies can further optimize the weight of these comorbidities in patients who underwent CABG to derive a CABG-specific comorbidity index score.

## Data Availability

All data produced in the present study are available upon reasonable request to the authors.

## Non-standard Abbreviations and Acronyms

AUC: area under curve
CABG: Coronary Artery Bypass Graft
CI: confidence interval
ECI: Elixhauser Comorbidity Index
ICD-10-CM: International Classification of Diseases, Tenth Revision, Clinical Modification
ICD-10-PCS: International Classification of Diseases, Tenth Revision, Procedure Coding System
IHD: ischemic heart disease
NIS: National (Nationwide) Inpatient Sample
ROC: receiver operating characteristic

## Acknowledgments

Renxi Li participated in all aspects of the study.

## Supplemental Material

Supplementary Table 1.

