## Supplementary Table 1 for "Predicting In-hospital Mortality Using Elixhauser Comorbidity in Patients Underwent Single and Multiple Coronary Artery Bypass Surgery"

### Supplemental Material

**Supplementary Table 1.** International Classification of Diseases, Tenth Revision, Procedure Coding System (ICD-10-PCS) codes used to extract patients who underwent 1, 2, 3, or 4+ Coronary Artery Bypass Surgery (CABG) in the National (Nationwide) Inpatient Sample (NIS) database.

| | **ICD-10-PCS** |
|---|---|
| **1 CABG** | 210083, 210088, 210089, 021008C, 021008W, 210093, 210098, 210099, 021009C, 021009F, 021009W, 02100A3, 02100A8, 02100A9, 02100AC, 02100AF, 02100AW, 02100J3, 02100J8, 02100J9, 02100JW, 02100K3, 02100K8, 02100K9, 02100KC, 02100KW, 02100Z3, 02100Z8, 02100Z9, 02100ZC, 02100ZF, 210488, 210489, 210493, 210498, 210499, 021049C, 021049W, 02104A3, 02104A8, 02104A9, 02104AC, 02104AF, 02104AW, 02104D4, 02104J9, 02104JW, 02104K3, 02104K9, 02104Z3, 02104Z8, 02104Z9, 02104ZC |
| **2 CABG** | 211083, 211089, 021108C, 021108W, 211093, 211098, 211099, 021109C, 021109F, 021109W, 02110A3, 02110A8, 02110A9, 02110AC, 02110AF, 02110AW, 02110J3, 02110J8, 02110J9, 02110JW, 02110K3, 02110K9, 02110KC, 02110KW, 02110Z3, 02110Z8, 02110Z9, 02110ZC, 02110ZF, 211489, 211493, 211498, 211499, 021149W, 02114A9, 02114AW, 02114JC, 02114KW, 02114Z3, 02114Z8, 02114Z9 |
| **3 CABG** | 021208W, 212093, 212098, 212099, 021209C, 021209F, 021209W, 02120A3, 02120A8, 02120A9, 02120AC, 02120AF, 02120AW, 02120J3, 02120J9, 02120JW, 02120K3, 02120K8, 02120K9, 02120KW, 02120Z3, 02120Z8, 02120Z9, 02120ZC, 02120ZF, 212493, 212499, 021249W, 02124A9, 02124AC, 02124AW, 02124D4, 02124Z3, 02124Z8, 02124Z9 |
| **4+ CABG** | 021208W, 212093, 212098, 212099, 021209C, 021209F, 021209W, 02120A3, 02120A8, 02120A9, 02120AC, 02120AF, 02120AW, 02120J3, 02120J9, 02120JW, 02120K3, 02120K8, 02120K9, 02120KW, 02120Z3, 02120Z8, 02120Z9, 02120ZC, 02120ZF, 212493, 212499, 021249W, 02124A9, 02124AC, 02124AW, 02124D4, 02124Z3, 02124Z8, 02124Z9 |
